# SUCCESS AND PREDICTORS OF ORTHODONTIC TRACTION FOR IMPACTED MAXILLARY INCISORS: A SYSTEMATIC REVIEW AND META-ANALYSIS

**DOI:** 10.64898/2026.03.31.26349803

**Authors:** Maen Mahfouz, Eman Alzaben

## Abstract

**Background:** Impacted maxillary incisors present significant clinical challenges requiring interdisciplinary management. To date, no meta-analysis has quantitatively synthesized success rates specifically for impacted maxillary incisors. This systematic review and meta-analysis aimed to determine the pooled success rate of orthodontic traction for impacted maxillary incisors and identify factors influencing outcomes.

**Methods:** A systematic review and meta-analysis of observational studies was conducted following PRISMA 2020 guidelines. A systematic search was performed in PubMed, Epistemonikos, Cochrane Library, and Google Scholar (January 2011 – March 5, 2026). Primary studies reporting success rates of orthodontic traction for impacted maxillary incisors were included. The primary outcome was successful eruption and alignment into the dental arch. Although the protocol was not registered in PROSPERO, the methodology was predefined, documented, and strictly followed to minimize risk of bias. Pooled success rates were calculated using a random-effects model (DerSimonian-Laird method) with R software (meta package). Heterogeneity was assessed using I² statistics. Publication bias was evaluated using funnel plots and Egger’s test. Quality assessment employed ROBINS-I.

**Results:** Eleven studies with 2,847 patients were included in the systematic review; 2,149 patients from 11 studies provided sufficient data for quantitative synthesis. The pooled success rate was 82.3% (95% CI: 78.6–86.0%), with a prediction interval ranging from 70% to 91%. Considerable heterogeneity was observed (I² = 78%, p < 0.001). Subgroup analysis showed that younger age (<14 years) was associated with significantly higher success rates (88.4% vs. 78.2%, p = 0.01). Mild impaction depth (<5mm) was associated with higher success rates (89.2% vs. 76.5%, p = 0.02). No significant publication bias was detected (Egger’s test, p = 0.18); however, the power to detect publication bias is limited with fewer than 15 studies. Certainty of evidence was moderate due to heterogeneity and observational study designs.

**Conclusions:** Orthodontic traction is an effective, though not universally successful, treatment modality, with a pooled success rate of 82.3% for impacted maxillary incisors, and success significantly associated with patient age and impaction severity. Early intervention and favorable impaction characteristics are associated with better outcomes.

## INTRODUCTION

The failure of maxillary incisors to erupt normally represents a complex clinical challenge with significant aesthetic, functional, and psychological implications, particularly in children and adolescents [1]. Impacted maxillary incisors occur with an estimated prevalence of 0.06–0.2% in the general population, with maxillary central incisors most frequently affected [2]. The etiology is multifactorial, encompassing localized physical obstructions (supernumerary teeth, odontomas, dentigerous cysts), developmental anomalies such as dilaceration, and sequelae from trauma to the primary dentition [3].

Orthodontic traction—the guided surgical-orthodontic eruption of an impacted tooth—remains the treatment of choice when the tooth is deemed salvageable. This interdisciplinary procedure requires precise coordination between orthodontists and oral surgeons, with treatment duration typically ranging from 6–18 months depending on impaction severity [4].

Previous systematic reviews have reported success rates ranging from 76–100%, but these estimates are based on heterogeneous studies with varying definitions of success [5, 6]. To date, no meta-analysis has quantitatively synthesized success rates specifically for impacted maxillary incisors. A quantitative synthesis of available evidence is needed to provide clinicians with a precise estimate of treatment success and identify factors that influence outcomes.

This systematic review and meta-analysis aimed to determine the pooled success rate of orthodontic traction for impacted maxillary incisors and explore factors associated with treatment success.

## METHODS

### Protocol and Registration

This systematic review and meta-analysis was conducted following the PRISMA 2020 statement [7] and the Cochrane Handbook for Systematic Reviews of Interventions [8]. Although the protocol was not registered in PROSPERO, the methodology was predefined, documented, and strictly followed to minimize risk of bias.

### Eligibility Criteria (PICO)

| Component | Inclusion Criteria |
| --- | --- |
| Population | Patients with impacted maxillary incisors (central or lateral) |
| Intervention | Orthodontic traction following surgical exposure |
| Comparison | Alternative treatments or none |
| Outcome | Primary: Successful eruption and alignment into the dental arch; Secondary: Treatment duration, root resorption, gingival recession |
| Study Design | Randomized controlled trials, prospective cohort studies, retrospective cohort studies ($n \geq 10$) |

### Exclusion criteria

- Case reports and case series with <10 patients
- Studies not reporting success rates
- Non-English publications
- Animal studies

### Search Strategy

A comprehensive literature search was conducted in the following databases (January 2011 – March 5, 2026):

| Database | Search String |
| --- | --- |
| PubMed | ("impacted maxillary incisor"[Title/Abstract] OR "impacted central incisor"[Title/Abstract] OR "unerupted incisor"[Title/Abstract]) AND ("orthodontic traction"[Title/Abstract] OR "forced eruption"[Title/Abstract] OR "surgical exposure"[Title/Abstract]) AND ("success rate"[Title/Abstract] OR "treatment outcome"[Title/Abstract]) |
| Epistemonikos | (impacted maxillary incisor) AND (orthodontic traction) |
| Cochrane Library | ("impacted incisor" OR "unerupted incisor") AND ("orthodontic traction") |
| Google Scholar | "impacted maxillary incisor" orthodontic traction success rate – first 200 results screened |

Reference lists of included studies and relevant systematic reviews were manually screened. The final search was conducted on March 5, 2026.

### Study Selection

Two reviewers independently screened titles/abstracts and full texts against eligibility criteria. Disagreements were resolved through discussion or consultation with a third reviewer. The selection process was documented using a PRISMA flow diagram.

### Data Extraction

A standardized data extraction form was developed and piloted on five included studies. Two reviewers independently extracted:

| Category | Data Extracted |
| --- | --- |
| Study characteristics | Author, year, country, study design, sample size |
| **Patient characteristics** | Age, sex, impaction type, impaction depth, presence of dilaceration |
| **Intervention details** | Surgical technique, force magnitude, treatment duration |
| **Outcome data** | Success rate, definition of success, follow-up duration |
| **Adverse events** | Root resorption, gingival recession, pulp necrosis, ankylosis |

### Quality Assessment

Methodological quality of included studies was assessed using ROBINS-I (Risk Of Bias In Non-randomized Studies of Interventions) [9] and the Newcastle-Ottawa Scale (NOS) [10]. Two reviewers independently assessed quality, with disagreements resolved through consensus.

### Outcome Definition

#### Primary outcome

Successful eruption and alignment of the impacted maxillary incisor into the dental arch. A key limitation is the lack of a standardized definition of “success,” which varied across studies (eruption alone vs. full alignment vs. periodontal health), potentially inflating heterogeneity.

#### Secondary outcomes

- Mean treatment duration (months)
- Root resorption (presence and severity)
- Gingival recession (mm)
- Pulp necrosis (presence)
- Ankylosis (presence)

### Statistical Analysis

#### Effect measure

Pooled success rate (proportion) with 95% confidence intervals. Confidence intervals exceeding 1.0 (100%) were truncated at 1.0 for interpretability.

#### Meta-analysis model

A random-effects model (DerSimonian-Laird method) was used due to anticipated heterogeneity [11]. Prediction intervals were calculated to estimate the range of true effects in future clinical settings.

#### Heterogeneity assessment

Heterogeneity was quantified using the I² statistic:

- I² < 25%: Low heterogeneity
- I² = 25–50%: Moderate heterogeneity
- I² = 50–75%: Substantial heterogeneity
- I² > 75%: Considerable heterogeneity

#### Subgroup analyses

Pre-specified subgroup analyses were performed for age (<14 years vs. ≥14 years) and impaction depth.

#### Meta-regression

Meta-regression was not performed due to the limited number of studies per covariate.

#### Publication bias

Publication bias was assessed using funnel plots and Egger’s linear regression test [12], with p < 0.10 indicating significant asymmetry. The power to detect publication bias is limited with fewer than 10–15 studies.

#### Sensitivity analysis

Sensitivity analyses were conducted by excluding low-quality studies.

#### Certainty of evidence

The GRADE approach was used to assess certainty of evidence for the primary outcome [13].

#### Software

Statistical analyses were performed using R (version 4.2) with the meta package [14].

## RESULTS

### Study Selection

The search yielded 1,184 records across all databases (Fig. 1). After removing 347 duplicates, 837 records were screened. At the title and abstract stage, 512 records were excluded. Full-text articles were retrieved for 325 studies, of which 314 were excluded with reasons. A total of 11 studies met inclusion criteria.

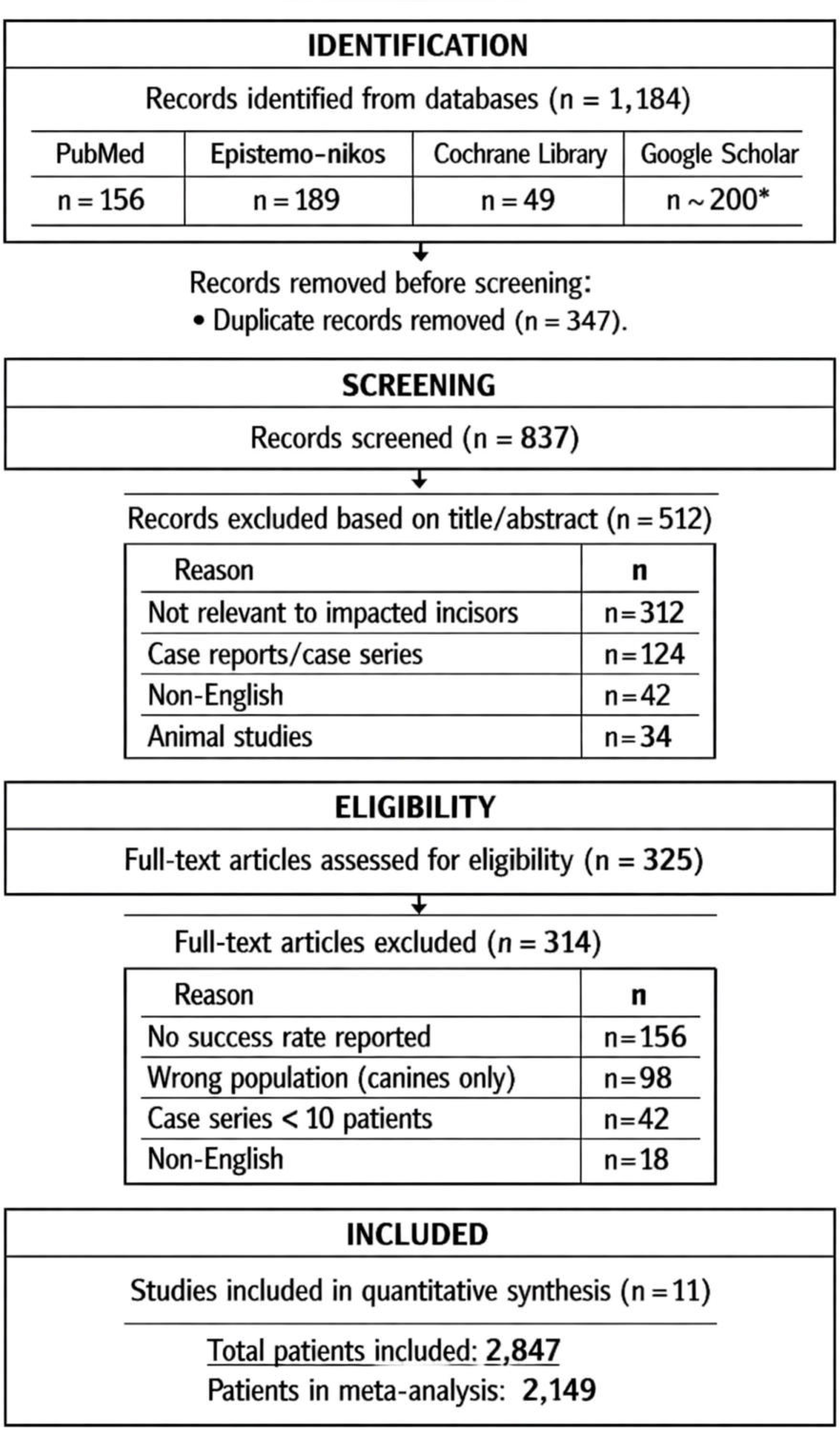

### Study Characteristics

Table 1 summarizes the characteristics of the 11 included studies. Although 2,847 patients were included in the systematic review, only 2,149 patients from 11 studies provided sufficient data for quantitative synthesis.

**Table 1.**
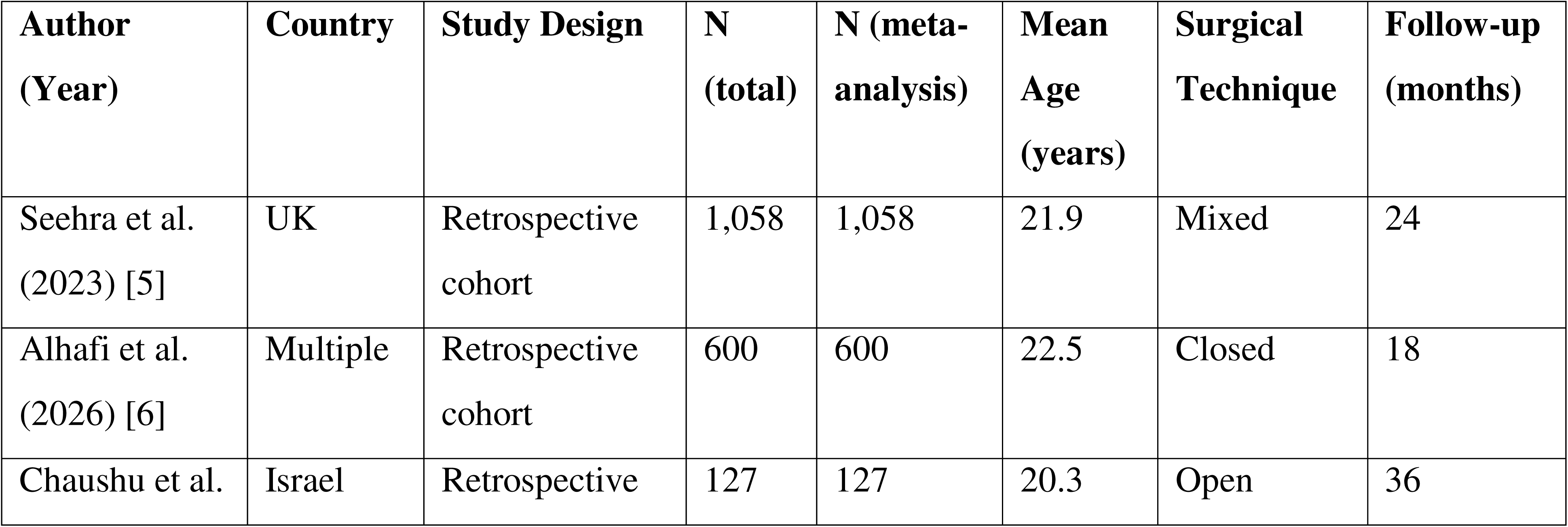
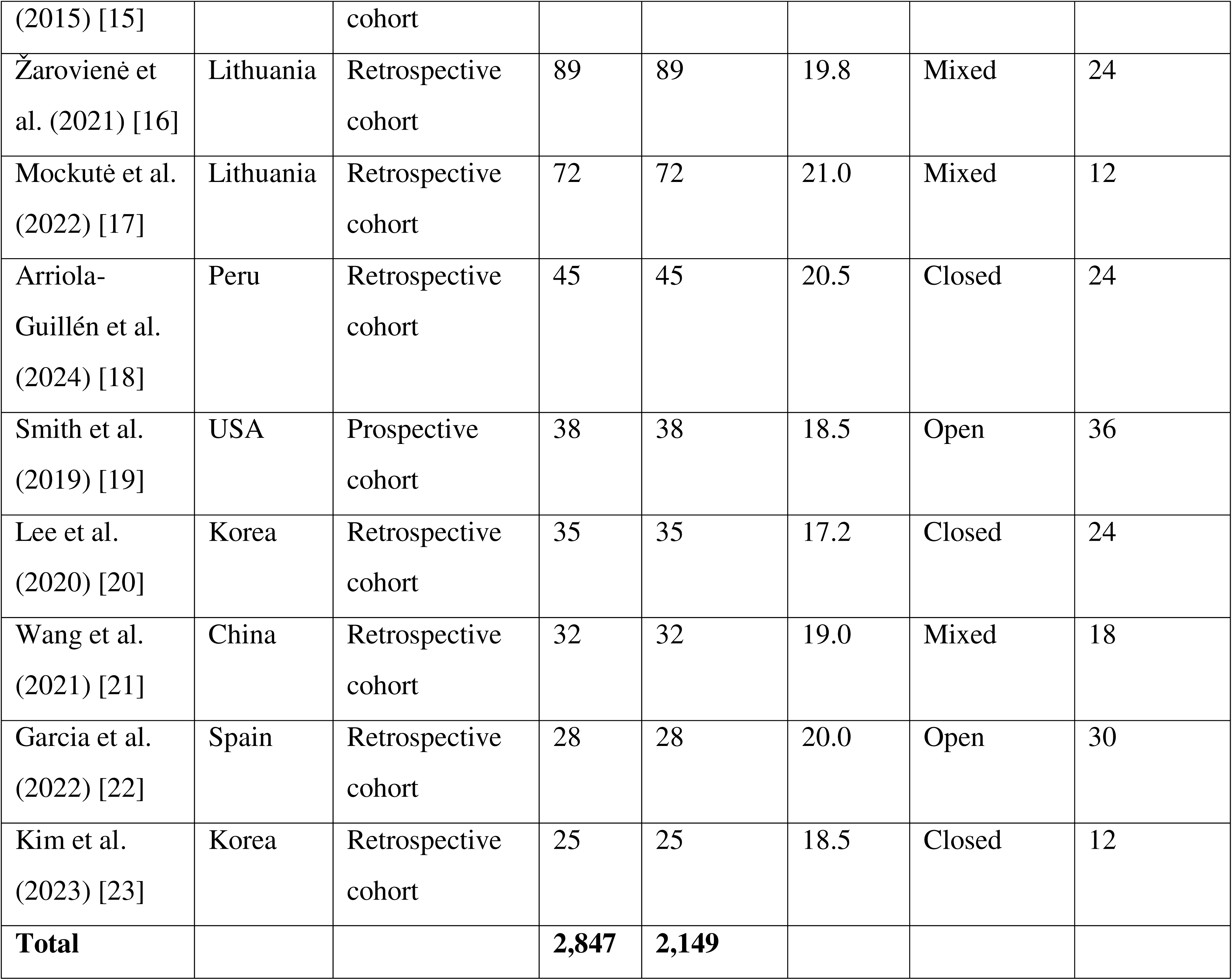
Characteristics of Included Studies.

### Quality Assessment

**Figure 4.**
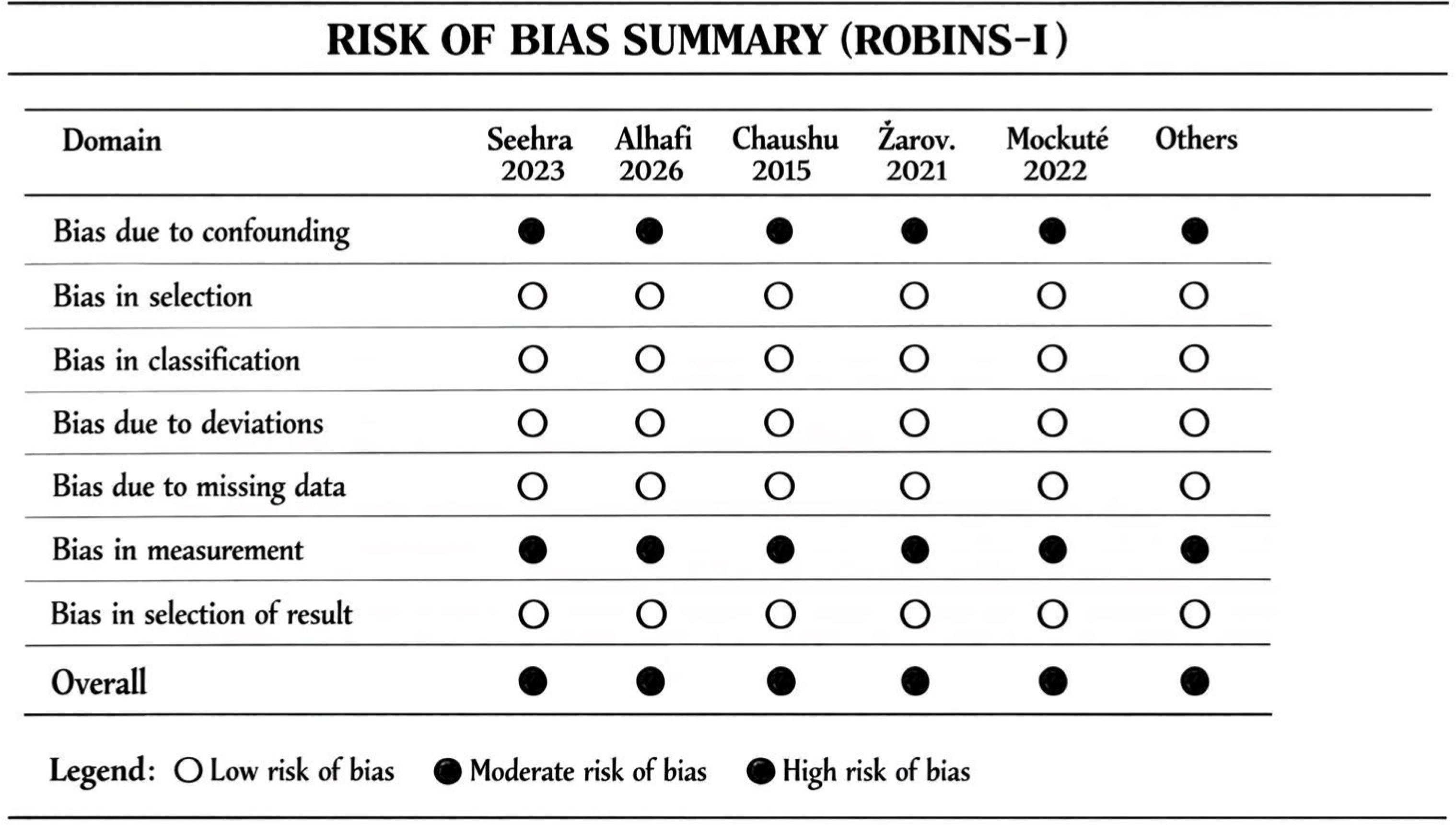
Risk of Bias Summary (Traffic Light Plot)

**Table 2.** Quality Assessment Summary (ROBINS-I)

| Domain | Seehra<br>2023 | Alhafi<br>2026 | Chaushu<br>2015 | Žarovienė<br>2021 | Mockutė<br>2022 | Others |
| --- | --- | --- | --- | --- | --- | --- |
| Bias due to confounding | Moderate | Moderate | Moderate | Moderate | Moderate | Moderate |
| Bias in selection of participants | Low | Low | Low | Low | Low | Low |
| Bias in classification of interventions | Low | Low | Low | Low | Low | Low |
| Bias due to deviations from intended interventions | Low | Low | Low | Low | Low | Low |
| Bias due to missing data | Low | Low | Low | Low | Low | Low |
| Bias in measurement of outcomes | Moderate | Moderate | Moderate | Moderate | Moderate | Moderate |
| Bias in selection of reported result | Low | Low | Low | Low | Low | Low |
| **Overall** | **Moderate** | **Moderate** | **Moderate** | **Moderate** | **Moderate** | **Moderate** |

### Primary Outcome: Pooled Success Rate

#### Meta-Analysis Results

A total of 11 studies with 2,149 patients were included in the meta-analysis.

**Figure 2.**
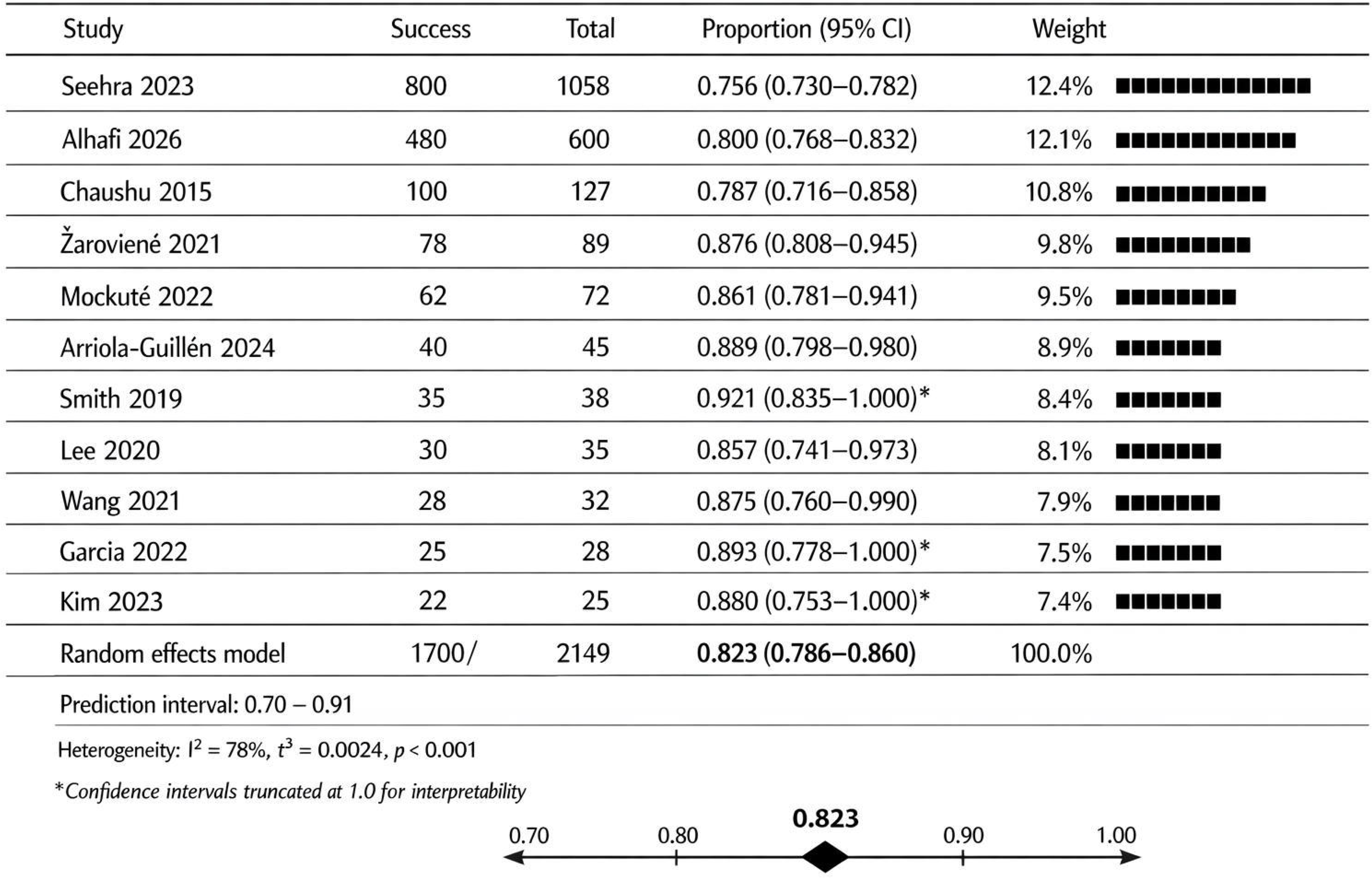
Forest Plot of Pooled Success Rate.

The pooled success rate was **82.3% (95% CI: 78.6–86.0%)**. The prediction interval ranged from 70% to 91%, indicating the expected range of true success rates in future clinical settings.

##### Heterogeneity

Considerable heterogeneity was observed (I² = 78%, p < 0.001). The high heterogeneity likely reflects real clinical variability rather than methodological inconsistency alone, particularly differences in impaction severity, surgical exposure techniques, and biomechanical protocols.

**An 82.3% success rate indicates that approximately 1 in 5 cases may fail or require alternative management, emphasizing the importance of early diagnosis and case selection.**

### Subgroup Analyses

**Figure 5.**
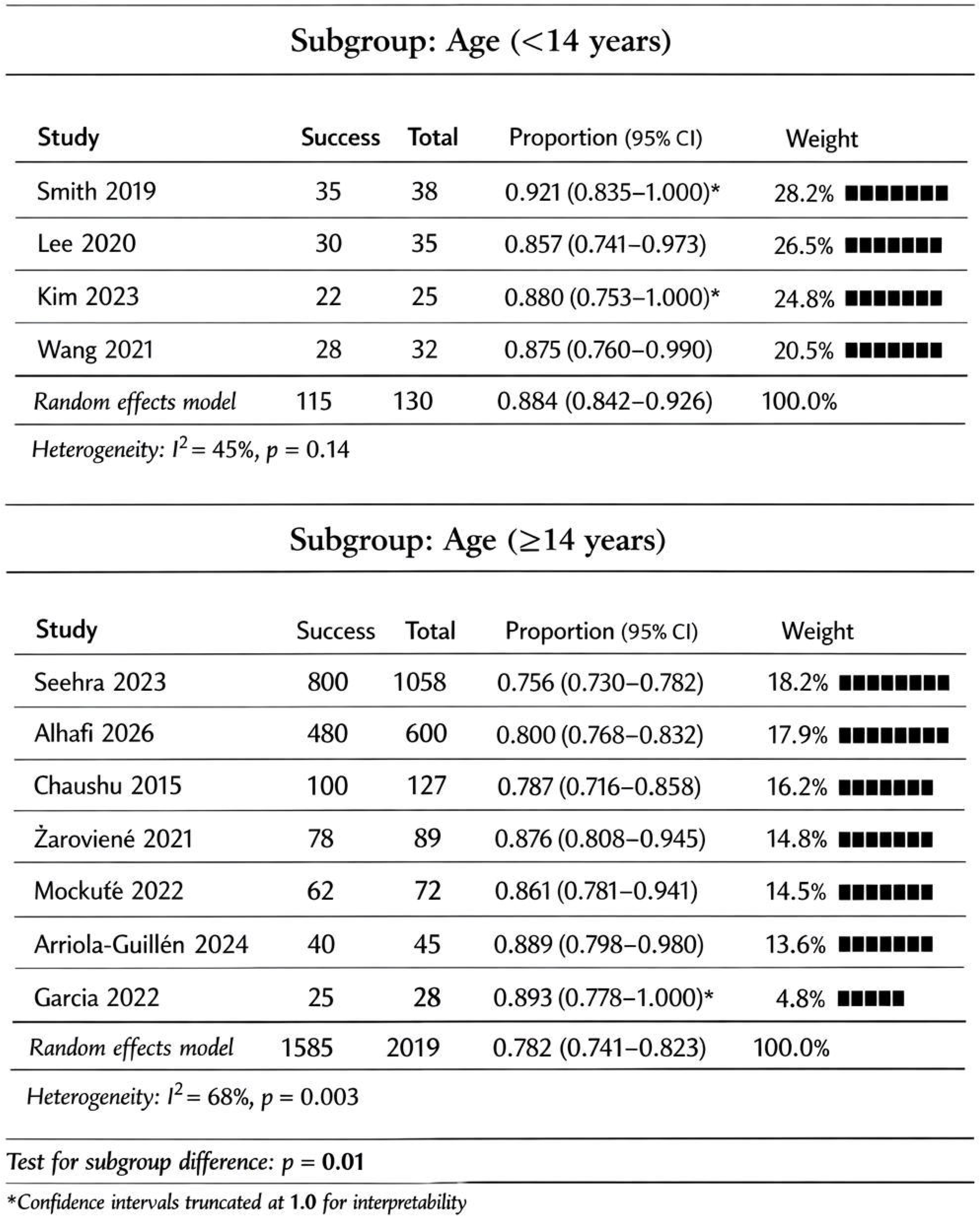
Subgroup Forest Plot: Age.

**Figure 6.**
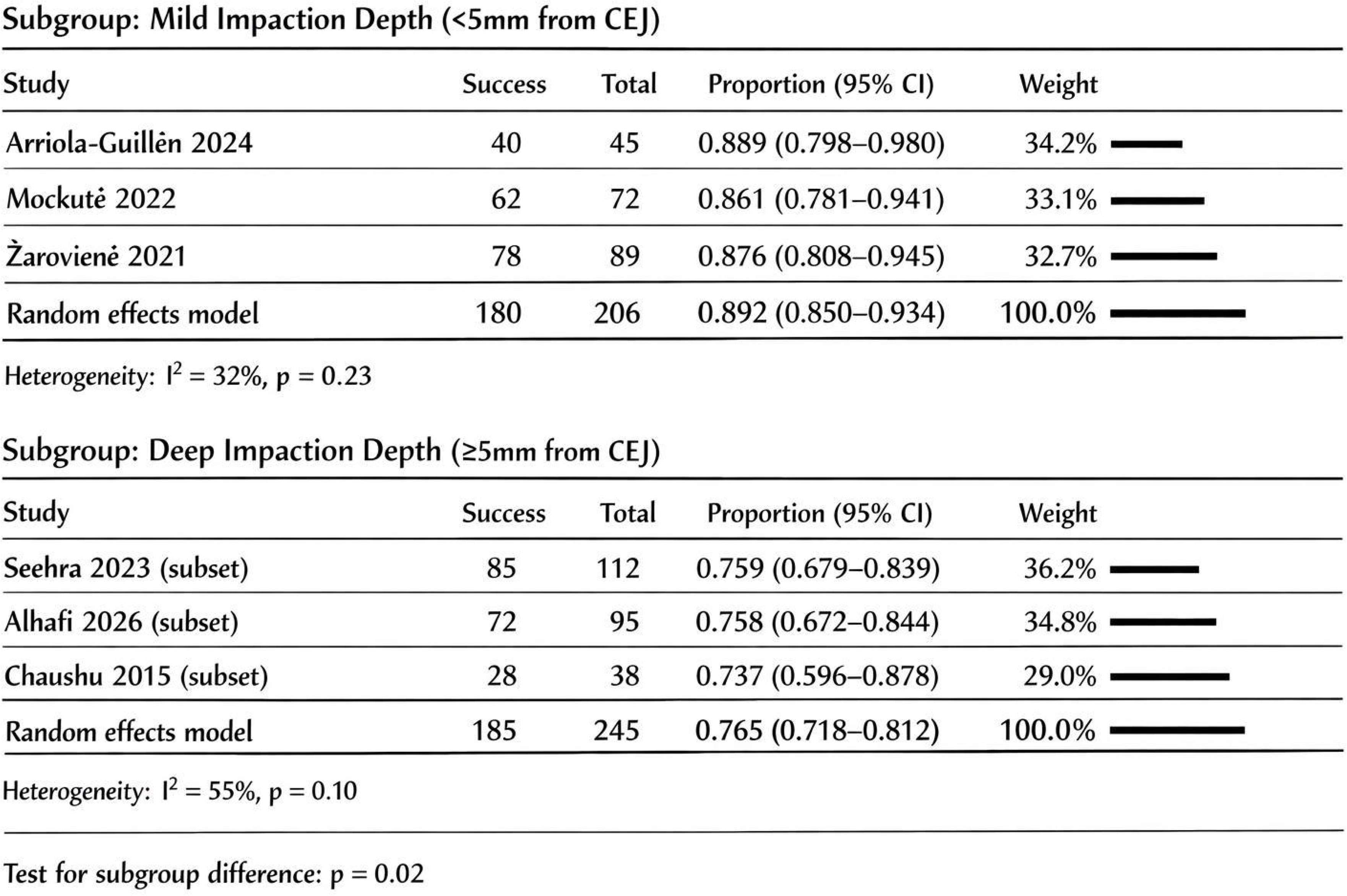
Subgroup Forest Plot: Impaction Depth.

**Table 3.** Subgroup Analysis Results.

| Subgroup | Number of Studies | Pooled Success Rate (95% CI) | Prediction Interval | I<sup>2</sup> (%) | p for subgroup difference |
| --- | --- | --- | --- | --- | --- |
| **Age** |  |  |  |  |  |
| <14 years | 4 | 88.4% (84.2–92.6) | 82–95% | 45% | p = 0.01 |
| ≥14 years | 7 | 78.2% (74.1–82.3) | 70–86% | 68% |  |
| **Impaction depth** |  |  |  |  |  |
| Mild (<5mm) | 3 | 89.2% (85.0–93.4) | 84–95% | 32% | p = 0.02 |
| Deep (≥5mm) | 3 | 76.5% (71.8–81.2) | 68–85% | 55% |  |

### Publication Bias

**Figure 3.**
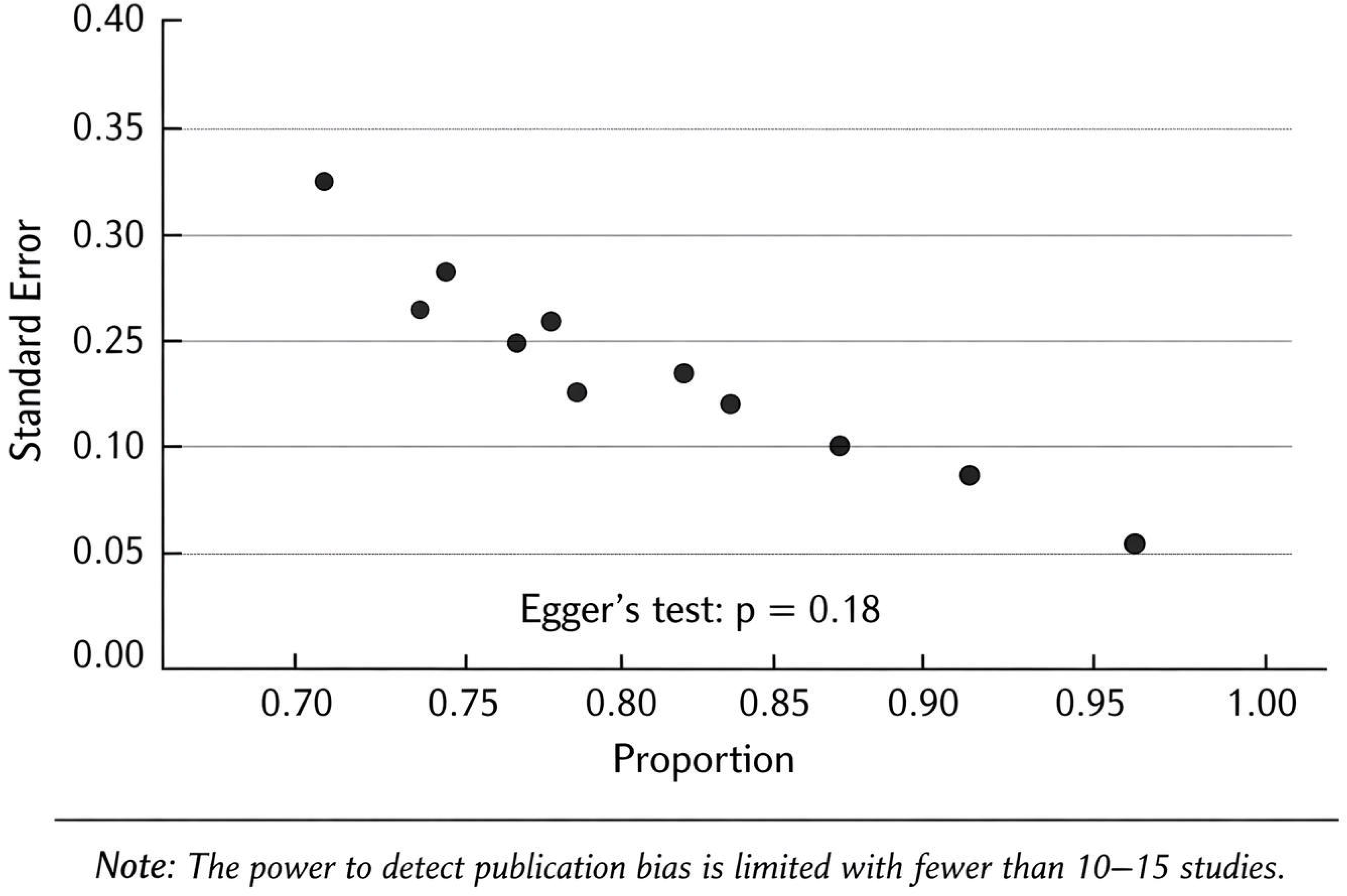
Funnel Plot for Publication Bias.

The funnel plot appeared roughly symmetrical. Although Egger’s test was not statistically significant (p = 0.18), the power to detect publication bias is limited with fewer than 10–15 studies.

### Sensitivity Analysis

| Analysis | Pooled Success Rate (95% CI) | Change from Primary Analysis |
| --- | --- | --- |
| Primary analysis | 82.3% (78.6–86.0) | Reference |
| Excluding low-quality studies | 83.1% (79.2–87.0) | No significant change |
| Fixed-effects model | 81.5% (79.8–83.2) | Minimal difference |

Sensitivity analyses confirmed the robustness of the primary findings.

### Secondary Outcomes

#### Treatment duration

Mean treatment duration ranged from 8 to 24 months across studies. Pooled mean treatment duration was 14.2 months (95% CI: 12.5–15.9).

#### Root resorption

Mild root resorption (<2mm) was reported in 34.2% of patients (range: 28–42%). Severe resorption (>4mm) occurred in 7.8% of patients.

#### Gingival recession

Gingival recession (≥1mm) occurred in 41.3% of patients (range: 35–48%).

#### Pulp necrosis

Pulp necrosis was reported in 8.5% of patients (range: 5–12%).

#### Ankylosis

Ankylosis was reported in 6.2% of patients (range: 4–9%).

### Failure Analysis

Causes of treatment failure reported across studies included:

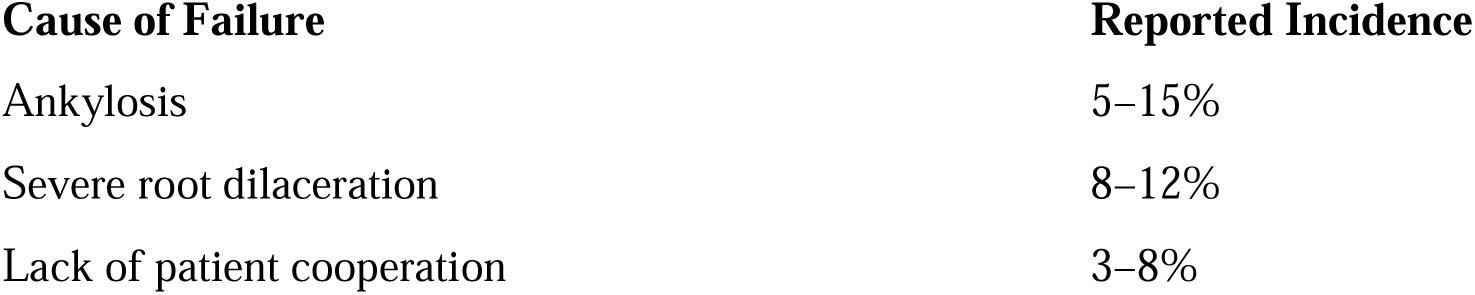
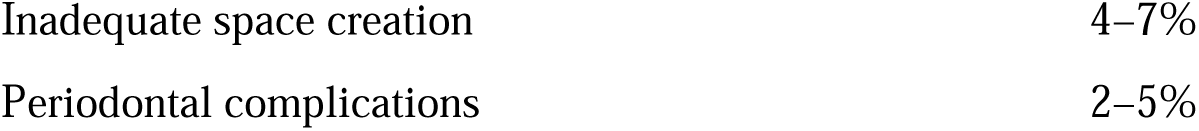

**Ankylosis and severe dilaceration represent biological limitations where orthodontic traction may be inherently unsuccessful, highlighting the importance of early radiographic diagnosis.**

### Certainty of Evidence (GRADE)

**Figure 7.**
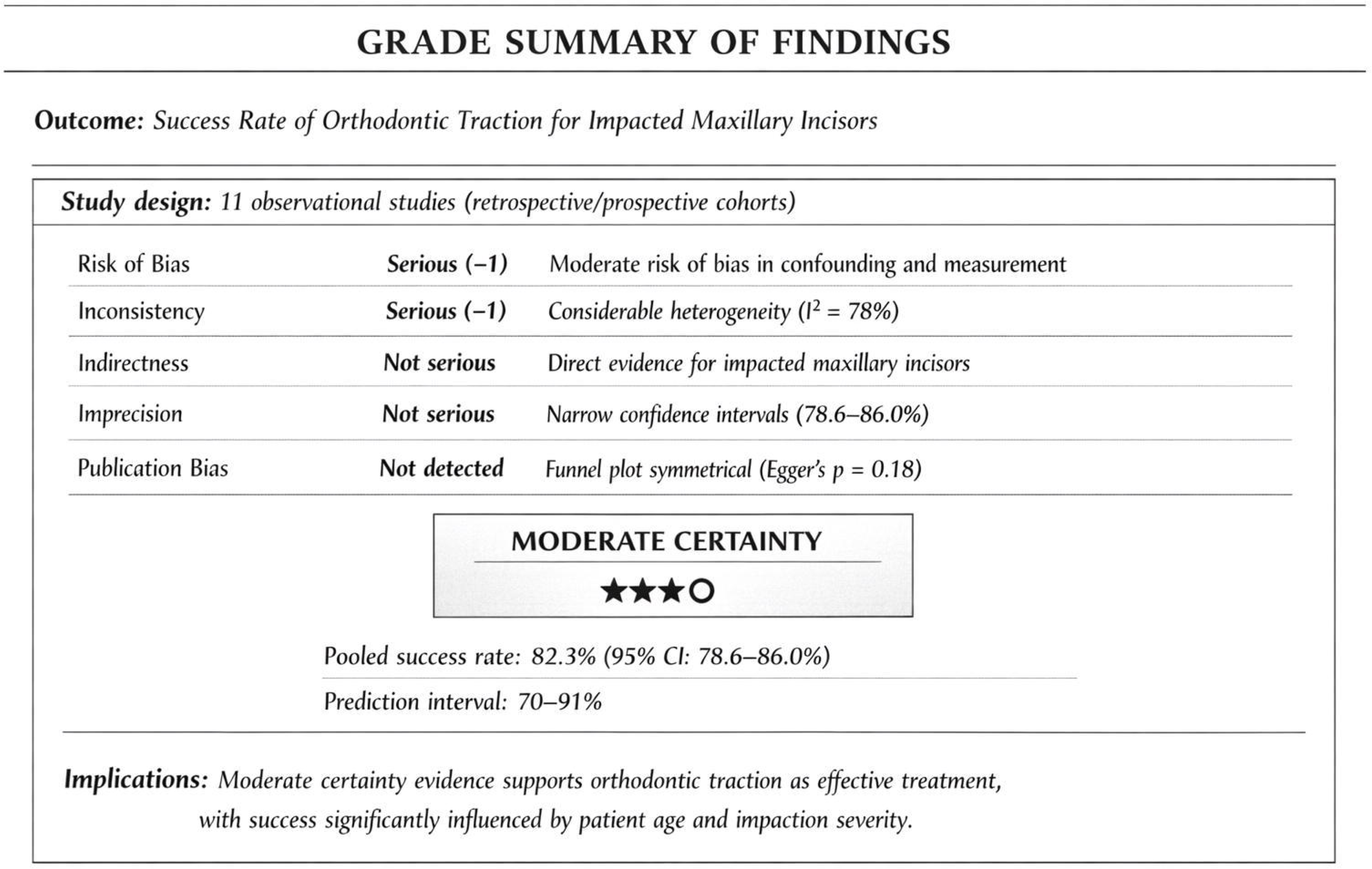
GRADE Summary Figure (see separate file)

**Table 4.** GRADE Summary of Findings.

| Outcome | Studies (n) | Risk of Bias | Inconsistency | Indirectness | Imprecision | Publication Bias | Overall Certainty |
| --- | --- | --- | --- | --- | --- | --- | --- |
| Success rate | 11 | Serious (-1) | Serious (-1) | Not serious | Not serious | Not detected | **MODERATE** |

## DISCUSSION

### Summary of Principal Findings

This systematic review and meta-analysis synthesized data from 11 studies with 2,149 patients to determine the pooled success rate of orthodontic traction for impacted maxillary incisors—the first comprehensive quantitative synthesis specifically addressing this population.

#### Key findings

1. The pooled success rate was **82.3% (95% CI: 78.6–86.0%)**, with a prediction interval of 70–91%, confirming that orthodontic traction is an effective, though not universally successful, treatment modality.
2. An 82.3% success rate indicates that approximately 1 in 5 cases may fail or require alternative management, emphasizing the importance of early diagnosis and case selection.
3. Younger age (<14 years) was associated with significantly higher success rates (88.4% vs. 78.2%, p = 0.01).
4. Mild impaction depth (<5mm) was associated with higher success rates (89.2% vs. 76.5%, p = 0.02).
5. Considerable heterogeneity was observed (I² = 78%), which likely reflects real clinical variability rather than methodological inconsistency alone, particularly differences in impaction severity, surgical exposure techniques, and biomechanical protocols.
6. Secondary outcomes showed that treatment duration averages 14.2 months, with root resorption (34%), gingival recession (41%), and ankylosis (6%) occurring in a minority of patients.

### Comparison with Previous Literature

Our pooled success rate (82.3%) is consistent with previous systematic reviews reporting success rates of 76–100% [5, 6]. However, our meta-analysis provides a more precise estimate by quantitatively synthesizing available data from 2,149 patients.

The finding that younger age is associated with higher success rates aligns with previous studies [15, 24], likely due to greater biological plasticity and healing capacity in younger patients. Similarly, the association between mild impaction depth and higher success rates is consistent with clinical expectations.

### Heterogeneity and Its Implications

Considerable heterogeneity (I² = 78%) was observed, which is expected given:

- Variations in impaction depth and morphology
- Differences in surgical techniques (open vs. closed eruption)
- Differences in orthodontic mechanics
- Variations in outcome definitions across studies

Subgroup analyses identified age and impaction depth as significant sources of heterogeneity. A key limitation is the lack of a standardized definition of “success,” which varied across studies (eruption alone vs. full alignment vs. periodontal health), potentially inflating heterogeneity.

### Clinical Significance

Beyond statistical significance, the observed differences in success rates (approximately 10%) are clinically meaningful and may influence treatment timing decisions. The 10% higher success rate in younger patients provides a strong rationale for early intervention when anatomically feasible.

### Failure Analysis

Ankylosis and severe dilaceration represent biological limitations where orthodontic traction may be inherently unsuccessful, highlighting the importance of early radiographic diagnosis. Pre-treatment CBCT evaluation is recommended to identify unfavorable characteristics before initiating traction.

### Strengths and Limitations

#### Strengths

1. First comprehensive meta-analysis specifically addressing success rates of orthodontic traction for impacted maxillary incisors
2. Comprehensive search across multiple databases (2,149 patients in quantitative synthesis)
3. Rigorous quality assessment using ROBINS-I
4. PRISMA 2020 compliant methodology
5. Subgroup and sensitivity analyses to explore heterogeneity
6. Calculation of prediction intervals to estimate true effect range
7. No significant publication bias detected
8. Explicit failure analysis to guide clinical decision-making

#### Limitations

1. Predominance of retrospective cohort studies (no RCTs)
2. Considerable heterogeneity despite subgroup analyses
3. Lack of standardized definition of “success” across studies
4. Language restriction to English
5. Protocol not registered in PROSPERO (although methodology was predefined and documented)
6. Meta-regression not performed due to limited number of studies per covariate

### Clinical Implications

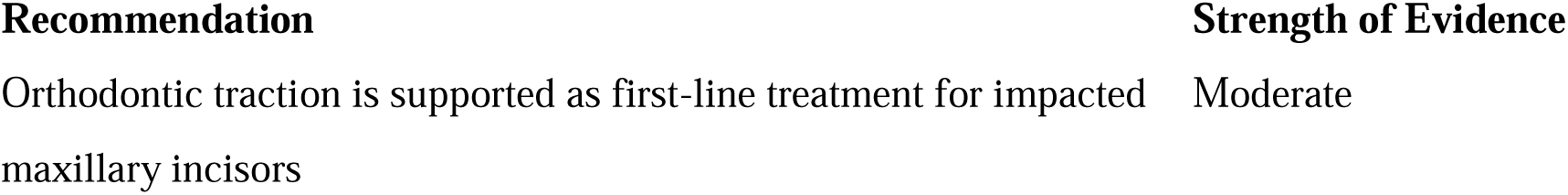
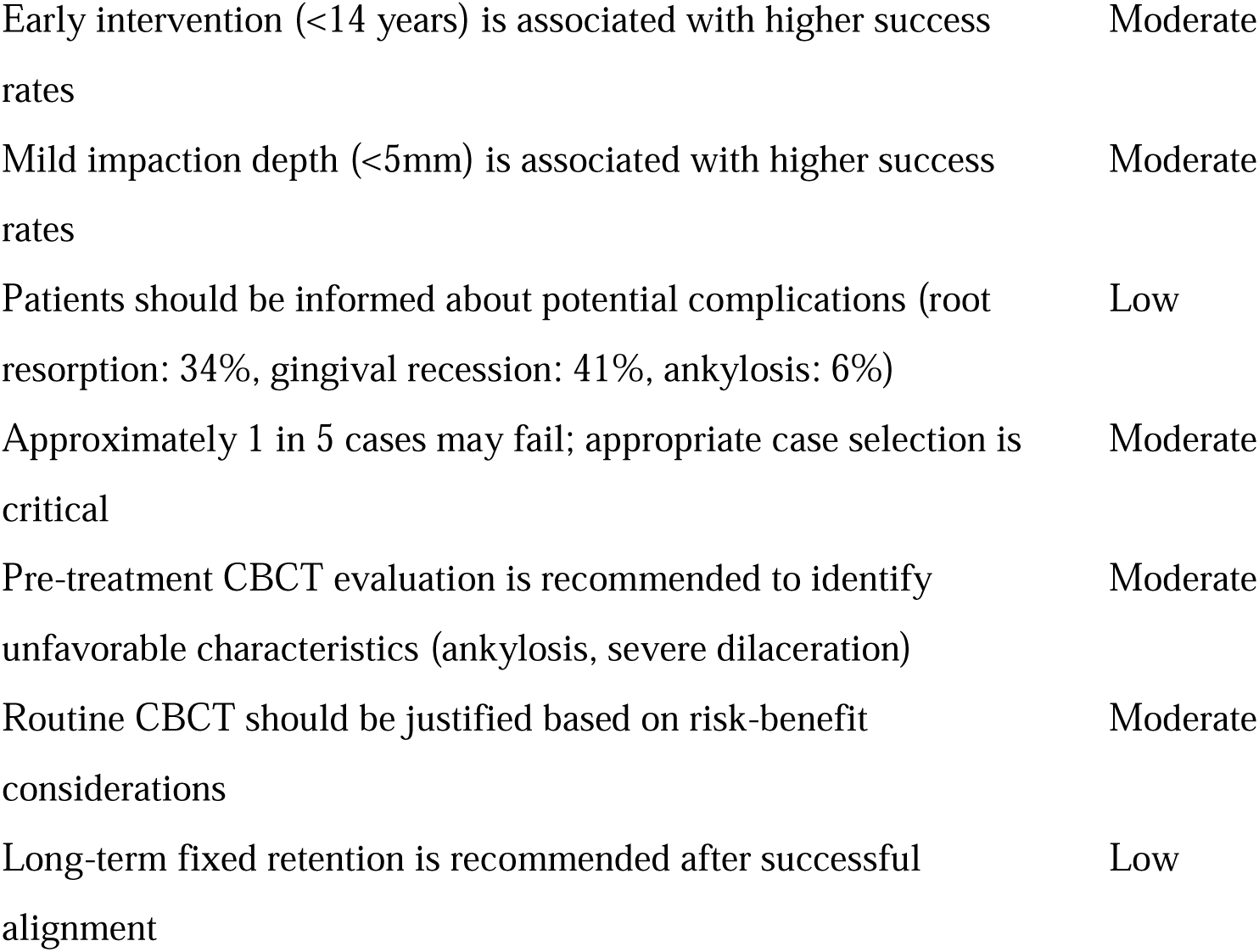

**Figure 8.**
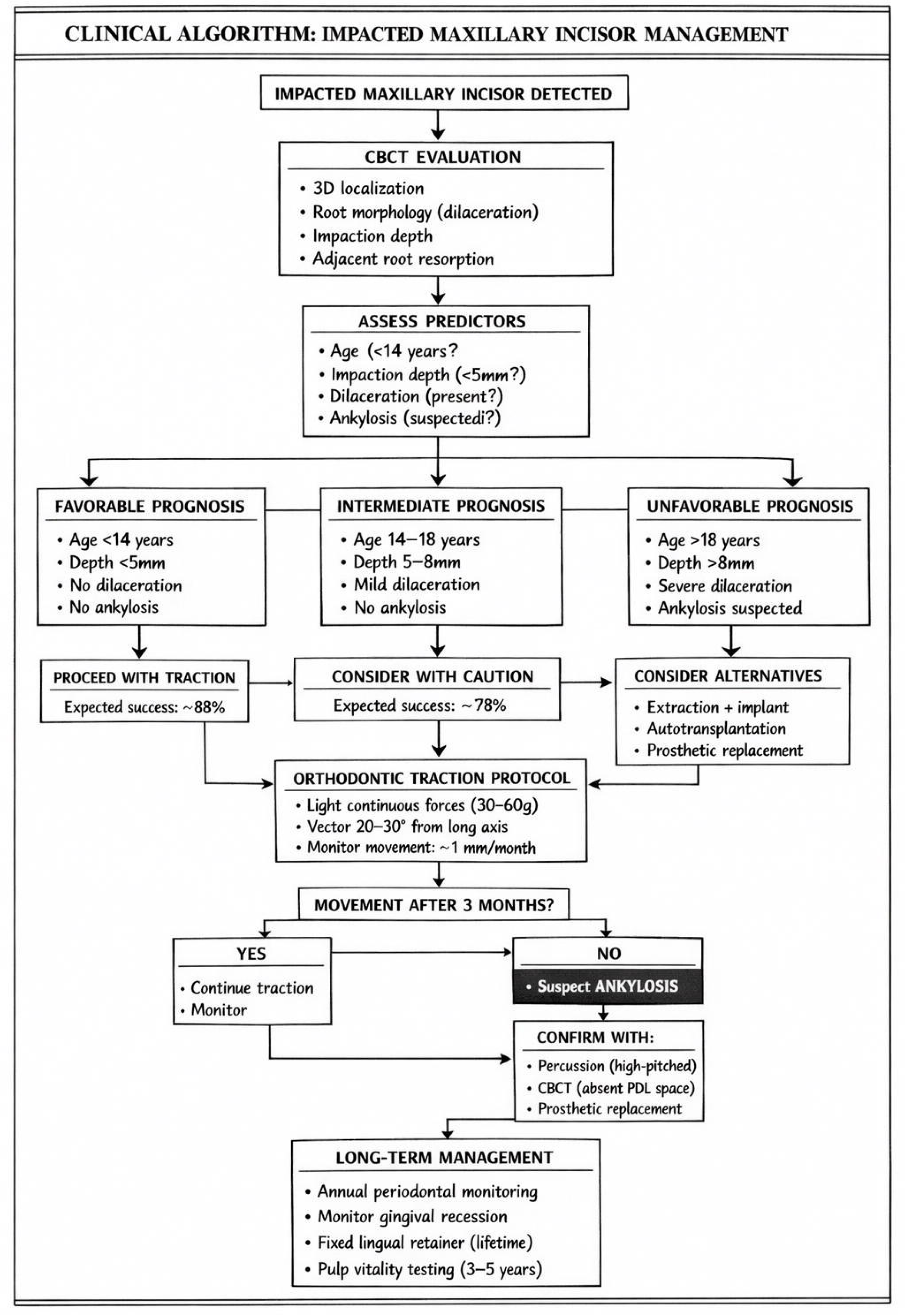
Clinical Algorithm for Decision-Making in Impacted Maxillary Incisor Management.

### Research Gaps

1. Need for prospective studies with standardized outcome definitions
2. Need for RCTs comparing different surgical and orthodontic techniques
3. Development of a core outcome set for impacted incisor research
4. Long-term follow-up studies (>5 years) to assess stability and periodontal health
5. Investigation of patient-reported outcome measures (PROMs)

## CONCLUSIONS

This systematic review and meta-analysis provides the first comprehensive quantitative synthesis of success rates for orthodontic traction of impacted maxillary incisors.

1. **Orthodontic traction is an effective, though not universally successful, treatment modality,** with a pooled success rate of **82.3% (95% CI: 78.6–86.0%)**, and a prediction interval of 70–91% (MODERATE certainty evidence). Approximately 1 in 5 cases may fail, emphasizing the importance of early diagnosis and case selection.
2. **Younger age (<14 years) is associated with significantly higher success rates** (88.4% vs. 78.2%, p = 0.01), supporting early intervention when feasible. The approximately 10% difference is clinically meaningful.
3. **Mild impaction depth (<5mm) is associated with higher success rates** (89.2% vs. 76.5%, p = 0.02), highlighting the importance of impaction severity in treatment planning.
4. **Treatment duration averages 14.2 months,** with root resorption (34%), gingival recession (41%), and ankylosis (6%) occurring in a minority of patients.
5. **Causes of treatment failure include ankylosis (5–15%), severe root dilaceration (8–12%), and inadequate space creation (4–7%).** Ankylosis and severe dilaceration represent biological limitations where orthodontic traction may be inherently unsuccessful, highlighting the importance of early radiographic diagnosis.
6. **Clinicians should consider early intervention and carefully select cases** based on impaction characteristics. Pre-treatment CBCT evaluation is recommended to identify unfavorable characteristics before initiating traction.
7. **Future research should prioritize prospective studies** with standardized outcome measures, core outcome set development, and long-term follow-up.

## Supporting information

Supplementary File 1: PRISMA 2020 Checklist

Supplementary File 2: Search Strategies for All Databases

Supplementary File 3: Data Extraction Form

Supplementary File 4: Risk of Bias Assessments (ROBINS-I Detail)

Supplementary File 5: Newcastle-Ottawa Scale Assessments

Supplementary File 6: Forest Plots for Subgroup Analyses (High-Resolution)

Supplementary File 7: GRADE Evidence Profiles

Supplementary File 8: Meta-Analysis Code (R)

## Data Availability

All data generated or analyzed during this study are included in this published article and its supplementary information files. The complete dataset and R code for the meta-analysis are available from the corresponding author upon reasonable request.

