## Supplementary File 1: PRISMA 2020 Checklist for "SUCCESS AND PREDICTORS OF ORTHODONTIC TRACTION FOR IMPACTED MAXILLARY INCISORS: A SYSTEMATIC REVIEW AND META-ANALYSIS"

| **Section/Topic** | **Item** | **Checklist Item** | **Reported on Page** |
| --- | --- | --- | --- |
| **TITLE** |  |  |  |
| Title | 1 | Identify the report as a systematic review. | Title page |
| **ABSTRACT** |  |  |  |
| Abstract | 2 | See PRISMA 2020 for Abstracts checklist. | Abstract |
| **INTRODUCTION** |  |  |  |
| Rationale | 3 | Describe the rationale for the review in the context of existing knowledge. | Introduction |
| Objectives | 4 | Provide an explicit statement of the objective(s) or question(s) the review addresses. | Introduction |
| **METHODS** |  |  |  |
| Eligibility criteria | 5 | Specify the inclusion and exclusion criteria for the review. | Methods |
| Information sources | 6 | Specify all databases, registers, websites, and other sources searched. | Methods |
| Search strategy | 7 | Present the full search strategies for all databases. | Methods |
| Selection process | 8 | Specify the methods used to decide whether a study met the inclusion criteria. | Methods |
| Data collection process | 9 | Specify the methods used to collect data from reports. | Methods |
| Data items | 10 | List and define all outcomes for which data were sought. | Methods |
| Study risk of bias assessment | 11 | Specify the methods used to assess risk of bias in the included studies. | Methods |
| Effect measures | 12 | Specify for each outcome the effect measure(s). | Methods |
| Synthesis methods | 13 | Describe the processes used to decide which studies were eligible for each synthesis. | Methods |
| Reporting bias assessment | 14 | Describe any methods used to assess risk of bias due to missing results. | Methods |
| Certainty assessment | 15 | Describe any methods used to assess certainty in the body of evidence. | Methods |
| **RESULTS** |  |  |  |
| Study selection | 16 | Describe the results of the search and selection process, ideally using a flow diagram. | Results, Figure 1 |
| Study characteristics | 17 | Cite each included study and present its characteristics. | Results, Table 1 |
| Risk of bias in studies | 18 | Present assessments of risk of bias for each included study. | Results, Table 2, Figure 4 |
| Results of individual studies | 19 | For all outcomes, present, for each study: summary statistics and effect estimates. | Results, Figure 2 |
| Results of syntheses | 20 | For each synthesis, briefly summarise the characteristics and risk of bias. | Results |
| Reporting biases | 21 | Present assessments of risk of bias due to missing results. | Results, Figure 3 |
| Certainty of evidence | 22 | Present assessments of certainty in the body of evidence. | Results, Table 4, Figure 7 |
| **DISCUSSION** |  |  |  |
| Discussion | 23 | Provide a general interpretation of the results in the context of other evidence. | Discussion |
| **OTHER INFORMATION** |  |  |  |
| Registration and protocol | 24 | Provide registration information for the review. | Methods |
| Support | 25 | Describe sources of financial or non-financial support. | Declarations |
| Competing interests | 26 | Declare any competing interests of review authors. | Declarations |
| Availability of data | 27 | Report which of the following are publicly available. | Declarations |
