## Supplementary File 2: Search Strategies for All Databases for "SUCCESS AND PREDICTORS OF ORTHODONTIC TRACTION FOR IMPACTED MAXILLARY INCISORS: A SYSTEMATIC REVIEW AND META-ANALYSIS"

**PubMed Search Strategy (January 2011 – March 5, 2026):**

("impacted maxillary incisor"[Title/Abstract] OR "impacted central incisor"[Title/Abstract] OR "unerupted incisor"[Title/Abstract]) AND ("orthodontic traction"[Title/Abstract] OR "forced eruption"[Title/Abstract] OR "surgical exposure"[Title/Abstract]) AND ("success rate"[Title/Abstract] OR "treatment outcome"[Title/Abstract])

**Epistemonikos Search Strategy:**

(impacted maxillary incisor) AND (orthodontic traction)

**Cochrane Library Search Strategy:**

("impacted incisor" OR "unerupted incisor") AND ("orthodontic traction")

**Google Scholar Search Strategy:**

"impacted maxillary incisor" orthodontic traction success rate

(first 200 results screened by relevance ranking)
