## Supplementary File 3: Data Extraction Form for "SUCCESS AND PREDICTORS OF ORTHODONTIC TRACTION FOR IMPACTED MAXILLARY INCISORS: A SYSTEMATIC REVIEW AND META-ANALYSIS"

| **Field** | **Description** |
| --- | --- |
| **Study ID** | Author, year |
| **Country** | Country where study was conducted |
| **Study Design** | RCT, prospective cohort, retrospective cohort |
| **Sample Size** | Number of patients |
| **Age** | Mean age (years) |
| **Sex** | Male/Female distribution |
| **Impaction Type** | Central incisor, lateral incisor |
| **Impaction Depth** | Mild (<5mm), Deep (≥5mm) |
| **Dilaceration** | Present/Absent |
| **Surgical Technique** | Open eruption, closed eruption, mixed |
| **Orthodontic Force** | Magnitude (g) |
| **Treatment Duration** | Months |
| **Success Rate** | Number successful / Total |
| **Definition of Success** | Eruption alone, full alignment, periodontal health |
| **Follow-up Duration** | Months |
| **Root Resorption** | Mild (<2mm), Severe (>4mm) |
| **Gingival Recession** | Yes/No, mm |
| **Pulp Necrosis** | Yes/No |
| **Ankylosis** | Yes/No |
