## Supplementary File 4: Risk of Bias Assessments (ROBINS-I Detail) for "SUCCESS AND PREDICTORS OF ORTHODONTIC TRACTION FOR IMPACTED MAXILLARY INCISORS: A SYSTEMATIC REVIEW AND META-ANALYSIS"

| **Study** | **Confounding** | **Selection** | **Classification** | **Deviations** | **Missing Data** | **Measurement** | **Reported Result** | **Overall** |
| --- | --- | --- | --- | --- | --- | --- | --- | --- |
| Seehra 2023 | Moderate | Low | Low | Low | Low | Moderate | Low | Moderate |
| Alhafi 2026 | Moderate | Low | Low | Low | Low | Moderate | Low | Moderate |
| Chaushu 2015 | Moderate | Low | Low | Low | Low | Moderate | Low | Moderate |
| Žarovienė 2021 | Moderate | Low | Low | Low | Low | Moderate | Low | Moderate |
| Mockutė 2022 | Moderate | Low | Low | Low | Low | Moderate | Low | Moderate |
| Arriola-Guillén 2024 | Moderate | Low | Low | Low | Low | Moderate | Low | Moderate |
| Smith 2019 | Moderate | Low | Low | Low | Low | Moderate | Low | Moderate |
| Lee 2020 | Moderate | Low | Low | Low | Low | Moderate | Low | Moderate |
| Wang 2021 | Moderate | Low | Low | Low | Low | Moderate | Low | Moderate |
| Garcia 2022 | Moderate | Low | Low | Low | Low | Moderate | Low | Moderate |
| Kim 2023 | Moderate | Low | Low | Low | Low | Moderate | Low | Moderate |
