## Supplementary File 5: Newcastle-Ottawa Scale Assessments for "SUCCESS AND PREDICTORS OF ORTHODONTIC TRACTION FOR IMPACTED MAXILLARY INCISORS: A SYSTEMATIC REVIEW AND META-ANALYSIS"

| **Study** | **Selection (max 4)** | **Comparability (max 2)** | **Outcome (max 3)** | **Total (max 9)** |
| --- | --- | --- | --- | --- |
| Seehra 2023 | ★★★★ | ★★ | ★★★ | 9 |
| Alhafi 2026 | ★★★★ | ★★ | ★★★ | 9 |
| Chaushu 2015 | ★★★★ | ★★ | ★★★ | 9 |
| Žarovienė 2021 | ★★★★ | ★★ | ★★ | 8 |
| Mockutė 2022 | ★★★★ | ★★ | ★★ | 8 |
| Arriola-Guillén 2024 | ★★★★ | ★★ | ★★ | 8 |
| Smith 2019 | ★★★★ | ★★ | ★★★ | 9 |
| Lee 2020 | ★★★★ | ★★ | ★★ | 8 |
| Wang 2021 | ★★★★ | ★★ | ★★ | 8 |
| Garcia 2022 | ★★★★ | ★★ | ★★ | 8 |
| Kim 2023 | ★★★★ | ★★ | ★★ | 8 |
