## Supplementary figures and images for "SUCCESS AND PREDICTORS OF ORTHODONTIC TRACTION FOR IMPACTED MAXILLARY INCISORS: A SYSTEMATIC REVIEW AND META-ANALYSIS"

### Supplementary File 6: Forest Plots for Subgroup Analyses (High-Resolution)

#### **Supplementary File 6: Forest Plots for Subgroup Analyses (High-Resolution**

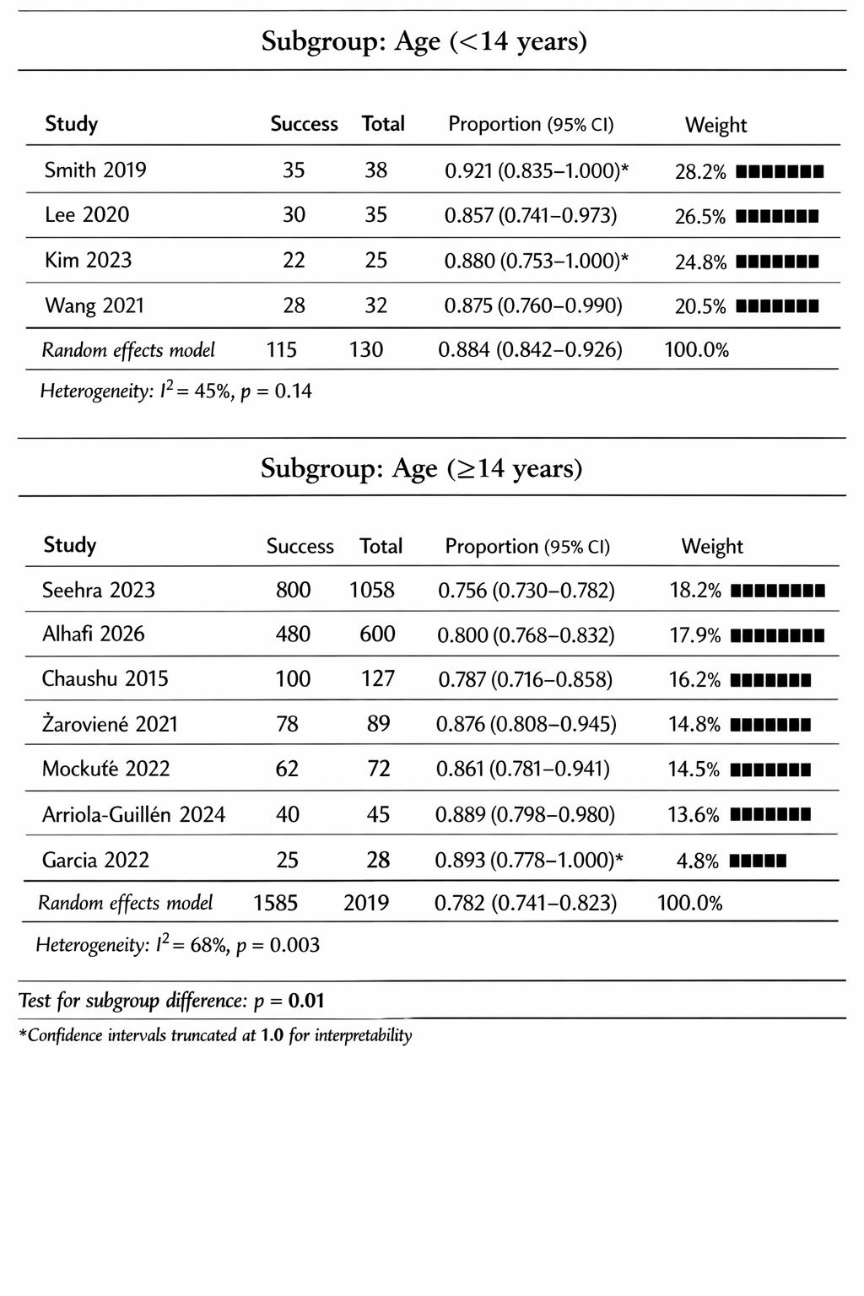

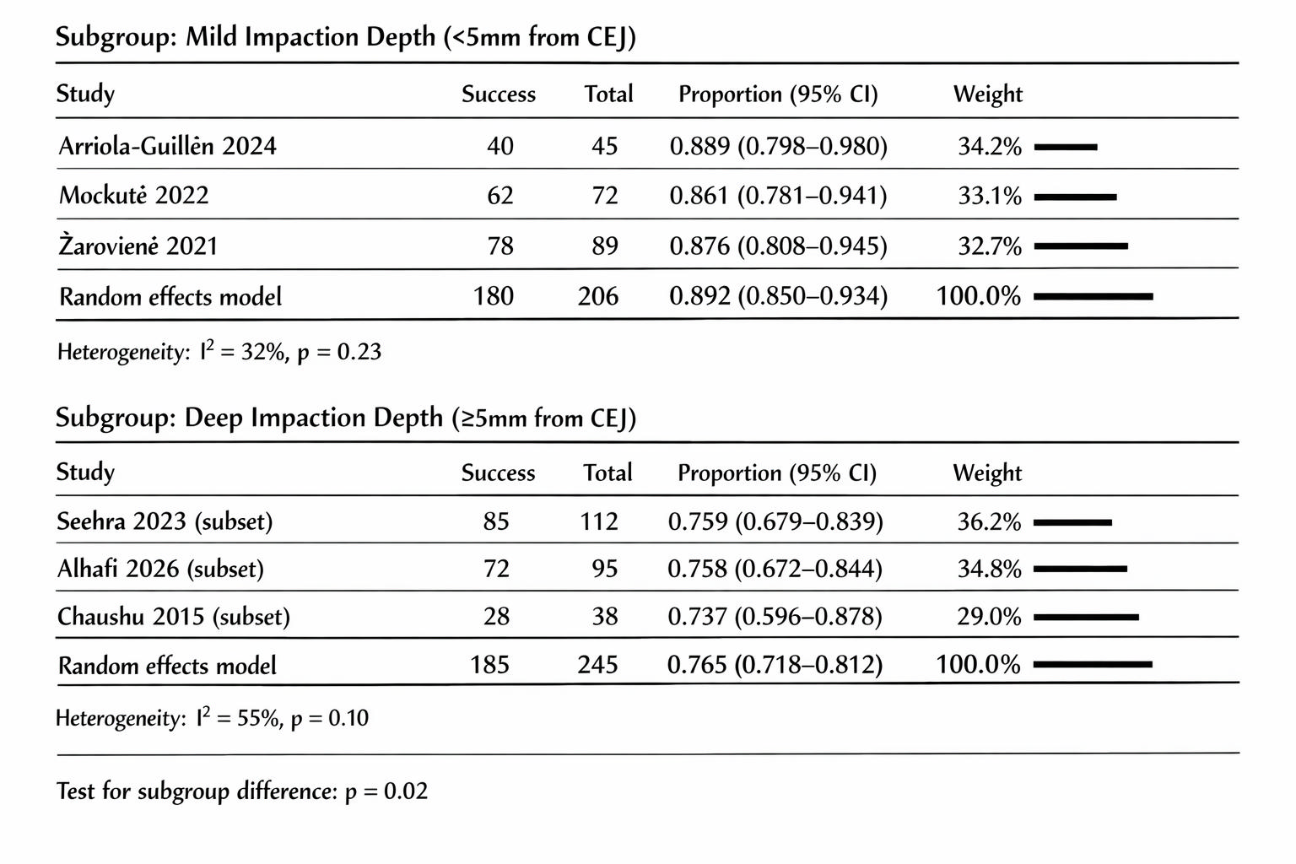
