## Supplementary File 7: GRADE Evidence Profiles for "SUCCESS AND PREDICTORS OF ORTHODONTIC TRACTION FOR IMPACTED MAXILLARY INCISORS: A SYSTEMATIC REVIEW AND META-ANALYSIS"

| **Quality Assessment** |  |  |  |  |  |  | **Summary of Findings** |  |
| --- | --- | --- | --- | --- | --- | --- | --- | --- |
| **No of studies** | **Study design** | **Risk of bias** | **Inconsistency** | **Indirectness** | **Imprecision** | **Publication bias** | **Overall certainty** | **Effect estimate (95% CI)** |
| 11 | Observational | Serious (-1) | Serious (-1) | Not serious | Not serious | Not detected | ⊕⊕⊕○ MODERATE | 82.3% (78.6–86.0%) |
