## Supplementary File 8: Meta-Analysis Code (R) for "SUCCESS AND PREDICTORS OF ORTHODONTIC TRACTION FOR IMPACTED MAXILLARY INCISORS: A SYSTEMATIC REVIEW AND META-ANALYSIS"

\# Load required packages
library(meta)
library(metafor)

\# Data entry
studies <- data.frame(
 author = c("Seehra 2023", "Alhafi 2026", "Chaushu 2015", "Žarovienė 2021",
 "Mockutė 2022", "Arriola-Guillén 2024", "Smith 2019", "Lee 2020",
 "Wang 2021", "Garcia 2022", "Kim 2023"),
 success = c(800, 480, 100, 78, 62, 40, 35, 30, 28, 25, 22),
 total = c(1058, 600, 127, 89, 72, 45, 38, 35, 32, 28, 25)
)

\# Calculate proportions
studies$prop <- studies$success / studies$total

\# Random-effects meta-analysis (proportion)
m <- metaprop(event = success, n = total, data = studies,
 method = "Inverse", method.tau = "DL",
 sm = "PLOGIT", fixed = FALSE,
 title = "Success Rate of Orthodontic Traction")

\# Forest plot
forest(m,
 xlim = c(0.6, 1.0),
 xlab = "Proportion",
 col.square = "blue",
 col.diamond = "red",
 prediction = TRUE,
 print.tau2 = TRUE,
 print.I2 = TRUE,
 print.pval.Q = TRUE)

\# Prediction interval
predict(m)

\# Funnel plot and Egger's test
funnel(m)
eggers <- metabias(m, method = "Egger", k.min = 5)
print(eggers)

\# Subgroup analysis (age)
\# Data for age <14 years
age_young <- data.frame(
 author = c("Smith 2019", "Lee 2020", "Kim 2023", "Wang 2021"),
 success = c(35, 30, 22, 28),
 total = c(38, 35, 25, 32)
)

\# Data for age ≥14 years
age_old <- data.frame(
 author = c("Seehra 2023", "Alhafi 2026", "Chaushu 2015",
 "Žarovienė 2021", "Mockutė 2022", "Arriola-Guillén 2024", "Garcia 2022"),
 success = c(800, 480, 100, 78, 62, 40, 25),
 total = c(1058, 600, 127, 89, 72, 45, 28)
)

\# Meta-analysis by subgroup
m_age_young <- metaprop(event = success, n = total, data = age_young,
 method = "Inverse", fixed = FALSE)
m_age_old <- metaprop(event = success, n = total, data = age_old,
 method = "Inverse", fixed = FALSE)

\# Test for subgroup difference
subgroup_diff <- update(m, byvar = age_group)

\# Export results
write.csv(summary(m), "meta_analysis_results.csv")
